# How the COVID-19 pandemic is favoring the adoption of digital technologies in healthcare: a literature review

**DOI:** 10.1101/2020.04.26.20080341

**Authors:** Davide Golinelli, Erik Boetto, Gherardo Carullo, Andrea Giovanni Nuzzolese, Maria Paola Landini, Maria Pia Fantini

## Abstract

**Background:** Healthcare is responding to the COVID-19 pandemic through the fast adoption of digital solutions and advanced technology tools. The aim of this study is to describe which digital solutions have been reported in the scientific literature and to investigate their potential impact in the fight against the COVID-19 pandemic.

**Methods:** We conducted a literature review searching PubMed and MedrXiv with terms considered adequate to find relevant literature on the use of digital technologies in response to COVID-19. We developed an impact score to evaluate the potential impact on COVID-19 pandemic of all the digital solutions addressed in the selected papers.

**Results:** The search identified 269 articles, of which 145 full-text articles were assessed and 124 included in the review after screening and impact evaluation. Of selected articles, most of them addressed the use of digital technologies for diagnosis, surveillance and prevention. We report that digital solutions and innovative technologies have mainly been proposed for the diagnosis of COVID-19. In particular, within the reviewed articles we identified numerous suggestions on the use of artificial-intelligence-powered tools for the diagnosis and screening of COVID-19. Digital technologies are useful also for prevention and surveillance measures, for example through contact-tracing apps or monitoring of internet searches and social media usage.

**Discussion:** It is worth taking advantage of the push given by the crisis, and mandatory to keep track of the digital solutions proposed today to implement tomorrow’s best practices and models of care, and to be ready for any new moments of emergency.

## 1. BACKGROUND

The Coronavirus Disease 2019 (COVID-19) pandemic, like all “serious disruptions” in human history, is causing an unprecedented health and economic crisis. At the same time though, this new situation is favoring the digital transition in many industries and in the society as a whole. This is the case, for example, of education.[1] The entire sector, from primary schools to Universities, has developed new strategies for teaching remotely, shifting from lectures in classrooms to live conferencing or online courses.[2]

Similarly, now - and perhaps more prominently in the forthcoming months - healthcare is responding to the COVID-19 pandemic through the fast adoption of digital solutions and advanced technology tools. In times of pandemic, digital technology can mitigate or even solve many challenges, thus improving health care delivery. This is currently being done to address acute needs that are a direct or indirect consequence of the pandemic (e.g. apps for patient tracing, remote triage emergency services, etc.). Nevertheless, many of the solutions that have been developed and implemented at the moment of the current emergency could consolidate in the near future, contributing to the definition and adoption of new digital-based models of care.

Although with a certain degree of digital divide, [3] the list of new digital solutions is rapidly growing. Beyond video-visits, these options include email and mobile-phone applications and can expand to include uses of wearable devices, chatbots, artificial-intelligence (AI) powered diagnostic tools, voice-interface systems, or mobile sensors such as smartwatches, oxygen monitors or thermometers. A new category of service is the oversight of persons in home quarantine and large-scale population surveillance. Telemedicine and remote consultation have already proven to be effective at a time when access to health services for non-COVID-19 or non-acute patients is prevented, impeded or postponed. In fact, according to Keesara et al., [4] rather than a model structured on the historically necessary model of in-person interactions between patients and their clinicians’ through a face-to-face model of care, today’s healthcare services and patient assistance can be guaranteed remotely through digital technologies.

Before the COVID-19 pandemic, it was expected that digital transformation in health care would have been as disruptive as that seen in other industries. However, as stated by Hermann et al.,[5] *“despite new technologies being constantly introduced, this change had yet to materialize*”.[6] It appears that now the spread of COVID-19 has finally provided an ineludible sound reason to fully embrace the digital transformation. Moreover, simulations show that many countries will probably face several waves of contagions and new lockdowns will probably occur. [7] Therefore it becomes necessary to review the digital technologies that have been used during the emergency period and possibly consider them for continued use over time or cyclically in the event of recurring outbreaks.

## 2. AIM OF THE STUDY

According to Hermann et al. [5] digital technologies can be categorized based on the healthcare needs they address: diagnosis, prevention, treatment, adherence, lifestyle, and patient engagement. We argue that it is necessary to understand which digital technologies have been adopted to face the COVID-19 crisis, and whether and how they can still be of any use after the emergency phase. In order to do this it is crucial to cover as many as possible aspects of digital technology use in healthcare in response to the COVID pandemic.

The aim of this study is therefore to describe which digital solutions have been reported in the early scientific literature and to investigate their potential impact in the fight against the COVID-19 pandemic.

## 3. METHODS

### 3.1. Literature search

We conducted a review of the scientific literature to include quantitative and qualitative studies using diverse designs to describe which digital solutions have been reported to respond and fight the COVID-19 pandemic. The initial search was implemented on May 4, 2020 and was limited to the time-span from January 1, 2020 to April 30, 2020. The search query consisted of terms considered by the authors to review the literature on the use of digital technologies in response to COVID-19. Therefore, we searched PubMed/MEDLINE using the following search terms and database-appropriate syntax:

> *(“COVID-19”[All Fields] OR “COVID-2019”[All Fields] OR “severe acute respiratory syndrome coronavirus 2”[Supplementary Concept] OR “severe acute respiratory syndrome coronavirus 2”[All Fields] OR “2019-nCoV”[All Fields] OR “SARS-CoV-2”[All Fields] OR “2019nCoV”[All Fields] OR ((“Wuhan”[All Fields] AND (“coronavirus”[MeSH Terms] OR “coronavirus”[All Fields])) AND (2019/12[PDAT] OR 2020[PDAT]))) AND (digital[Title/Abstract] OR technology[Title/Abstract])]*

We also searched MedrXiv/BiorXiv^1^ (a preprint server for health science paper) section COVID-19/SARS-CoV-2 for digital technologies-related studies with the following search string: *COVID-19 digital technology*, with the same time-span restriction applied to the PubMed search.

We placed a language restriction for English, without other limits. A two-stage screening process was used to assess the relevance of identified studies. For the first level of screening, only the title and abstract were reviewed to preclude waste of resources in procuring articles that did not meet the minimum inclusion criteria. For the second level of screening all citations deemed relevant after title and abstract screening were procured for subsequent review of the full-text article.

### 3.2. Impact assessment

A form was developed to extract article characteristics. We categorized the papers retrieved according to the healthcare needs addressed (diagnosis, prevention, treatment, adherence, lifestyle, patient engagement and other). The definition of each healthcare need is reported in Table 1. We added “surveillance” as an additional healthcare need,[5] given the importance of early identification and confinement of COVID-19 patients, and a category “other” to include any further category not considered.

**Table 1.** Definition of the healthcare needs addressed by digital technologies.

| Use of technology | Definition |
| --- | --- |
| **Diagnosis** | The process of determining which disease or condition explains a person's symptoms and signs. |
| **Surveillance** | The continuous, systematic collection, analysis and interpretation of health-related data needed for the planning, implementation, and evaluation of public health practice |
| **Prevention** | Preventing the occurrence of a disease (e.g. by reducing risk factors) or by halting a disease and averting resulting complications after its onset. |
| **Adherence** | The degree to which a patient correctly follows medical advice. |
| **Treatment** | The use of an agent, procedure, or regimen, such as a drug, surgery, or exercise, in an attempt to cure or mitigate a disease |
| **Lifestyle** | Adoption and sustaining behaviors that can improve health and quality of life |
| **Patient engagement** | To actively involve people in their health and health care |

To evaluate the potential impact on COVID-19 pandemic of all the digital solutions addressed in the selected papers, we developed an impact score, which defines the impact on various healthcare targets, the degree of innovation and the scalability to other geographical areas. The score is calculated summing the following items’ score related to the characteristics of the digital solution proposed by each article:

- Healthcare system targets: score ranging from 1 to 4 depending on how many areas of the digital health services are addressed by a particular digital solution. We chose four target areas according to the Expert Panel on Effective Ways of Investigating in Health (EXPH) of the European Commission classification of digital health services [8]: (i) interventions for clients; (ii) for healthcare providers; (iii) for health system; (iv) for data services.
- Grade of innovation: from 1 to 4 depending on the degree of innovation (Supporting=1; Complementing=2; Substituting=3; Innovating=4). We have categorized the digital health solutions proposed in each paper according to the EXPH classification on the final use of digital tools [8].
- Scalability to other geographical areas: from 0 to 2 depending on the possibility to “transfer” the proposed digital solution into other regions or countries (Not possible=0; Possible, but only for limited regions/countries=1; Possible, globally=2).

The total score defines the overall potential impact of a particular digital solution on the COVID-19 pandemic, ranging from a minimum of 2 to a maximum of 10 (a solution that addresses all four areas of the digital health services, with a high degree of innovation and that is possible to implement globally). All articles with a score ≥ 5 have been deemed relevant in terms of degree of innovation, potential to impact on the COVID-19 pandemic outcome, and scalability to other geographical areas, and therefore included in the study (Figure 1). We then built a scoring rubric by cross-classifying the use of technology with the type of technology.

Lastly, in order to determine the attention received by the scientific community and the general public for each paper included in the study we traced altmetrics measures. Altmetrics, meant as a subset of scientometrics,[9] is the study and use of scholarly impact measures based on activity in online tools and environments. In order to consider the research impact of each article, we relied on the Computed Impact Score (CIS), which we have calculated as reported in previous papers.[10, 11] All papers with a CIS score above the 90% quantile threshold have been deemed valuable and listed in the results.

## 4. RESULTS

The search identified 269 articles (174 from Pubmed, 95 from MedrXiv), of which 145 full-text articles were assessed and 124 included in the review after screening and impact assessment (Figure 1).

Of selected articles, 65 (52.4%) addressed the use of digital technologies for diagnosis (Table 2), 46 (37.1%) for surveillance, 46 (37.1%) for prevention, 38 (30.6%) for treatment 15 (12.1%) for adherence, 12 (9.7%) for lifestyle, 11 (8.9%) for patient engagement and 6 (4.8%) for other purposes.

**Table 2.** Categories of healthcare needs addressed by digital technologies in the articles selected. N: absolute number of times healthcare needs are addressed.

| Healthcare need | N | %* |
| --- | --- | --- |
| Diagnosis | 65 | 52.4 |
| Surveillance | 46 | 37.1 |
| Prevention | 46 | 37.1 |
| Treatment | 38 | 30.6 |
| Adherence | 15 | 12.1 |
| Lifestyle | 12 | 9.7 |
| Patient engagement | 11 | 8.9 |
| Other | 6 | 4.8 |

\*The total is higher than 100% since some articles may include technologies used to address more than one healthcare need.

The impact assessment allowed us to select and include articles about digital technologies deemed impactful on the COVID-19 pandemic in terms of degree of innovation, potential impact on various healthcare system areas and scalability to other geographical areas (Table 3).

**Table 3.** Articles included in the literature review. Here reported the main characteristics and impact assessment of each paper analyzed.

| Reference | Area of health services addressed | Impact on healthcare services | Scalability to other geographical areas | Potential digital innovation Impact score | Reference | Area of health services addressed | Impact on healthcare services | Scalability to other geographical areas | Potential digital innovation Impact score |
| --- | --- | --- | --- | --- | --- | --- | --- | --- | --- |
| 12 | C, HP, HS, D | Innovating | Global | 10 | 72 | HP | Complementing | Global | 5 |
| 13 | C, HP, HS, D | Innovating | Local | 9 | 73 | HP | Complementing | Global | 5 |
| 14 | C, HP, HS, D | Innovating | Local | 9 | 74 | C, HP, HS, D | Innovating | Global | 10 |
| 15 | C, HP | Supporting | Global | 5 | 75 | HS, D | Innovating | Local | 7 |
| 16 | C, HS, D | Innovating | Local | 8 | 76 | C, HP | Innovating | Local | 7 |
| 17 | HP | Complementing | Global | 5 | 77 | C, HP | Innovating | Local | 7 |
| 18 | HP | Complementing | Global | 5 | 78 | C, HS, D | Innovating | Local | 8 |
| 19 | C, HP, HS | Innovating | Global | 9 | 79 | C, HP, HS, D | Innovating | Local | 9 |
| 20 | HP | Complementing | Global | 5 | 80 | C, HP | Innovating | Global | 8 |
| 21 | HP | Complementing | Global | 5 | 81 | C, HP, HS, D | Innovating | Local | 9 |
| 22 | HP, HS | Supporting | Global | 5 | 82 | C, HP | Innovating | Local | 7 |
| 23 | HP | Complementing | Global | 5 | 83 | C, HP | Innovating | Local | 7 |
| 24 | C, HS, D | Innovating | Local | 8 | 84 | C, HP | Innovating | Local | 7 |
| 25 | C, HP, HS, D | Innovating | Global | 10 | 85 | C, HO | Innovating | Global | 8 |
| 26 | C, HS, D | Innovating | Local | 8 | 86 | HP, HS, D | Supporting | Global | 6 |
| 27 | HP | Complementing | Global | 5 | 87 | C, HP, HS, D | Supporting | No [0] | 5 |
| 28 | HP | Complementing | Global | 5 | 88 | C, HP, HS, D | Innovating | Global | 10 |
| 29 | C, HP, HS, D | Innovating | Local | 9 | 89 | HP, D | Complementing | Global | 6 |
| 30 | C, HP, HS, D | Innovating | Global | 10 | 90 | HS, D | Supporting | Global | 5 |
| 31 | C, HP | Innovating | Global | 6 | 91 | C, HP, | Supporting | Global | 5 |
| 32 | HP | Complementing | Global | 5 | 92 | C, HP | Innovating | Global | 8 |
| 33 | C, HS, D | Innovating | Local | 8 | 93 | C, HP | Innovating | Global | 8 |
| 34 | C, HP | Innovating | Local | 7 | 94 | C, HP, HS, D | Innovating | Global | 10 |
| 35 | C, HS | Innovating | Local | 7 | 95 | C, HP, HS, D | Complementing | Global | 8 |
| 36 | C, HP, HS, D | Innovating | Local | 9 | 96 | HP, HS, D | Complementing | Global | 7 |
| 37 | C, HS, D | Innovating | Local | 8 | 97 | HP; HS | Innovating | Global | 8 |
| 38 | C, HP | Innovating | Global | 8 | 98 | C, HP, HS, D | Innovating | Global | 10 |
| 4 | C, HP, HS, D | Innovating | Local | 9 | 99 | C, HP, HS, D | Innovating | Global | 10 |
| 39 | C, HS, D | Innovating | Local | 8 | 100 | C, HP, HS, D | Innovating | Local | 9 |
| 40 | C, HP | Supporting | Global | 5 | 101 | C, HP | Complementing | Global | 6 |
| 41 | C, HS, D | Innovating | Local | 8 | 102 | C, HP | Complementing | Global | 6 |
| 42 | HS, D | Innovating | Global | 8 | 103 | D | Innovating | Global | 7 |
| 43 | C, HS, D | Innovating | Local | 8 | 104 | C, HP, HS, D | Supporting | Global | 7 |
| 44 | C, HS, D | Innovating | Local | 8 | 105 | C, HP, HS, D | Supporting | Global | 7 |
| 45 | HP | Complementing | Global | 5 | 106 | HS, D | Innovating | Global | 8 |
| 46 | C, HP, HS, D | Innovating | Local | 9 | 107 | C, HS | Supporting | Global | 5 |
| 47 | C, HP, HS, D | Innovating | Local | 9 | 108 | C, HP, HS, D | Substituting | Global | 9 |
| 48 | C, HS, D | Innovating | Local | 8 | 109 | C | Substituting | Global | 5 |
| 49 | HS, D | Complementing | Local | 5 | 110 | C, HP, HS | Complementing | Global | 7 |
| 50 | HS, D | Innovating | Global | 8 | 111 | C, HP, D | Innovating | Global | 9 |
| 51 | C, HS, D | Innovating | Local | 8 | 112 | C, HP, HS | Supporting | Local | 5 |
| 52 | C, HP | Complementing | Global | 6 | 113 | C, HP, HS, D | Complementing | Global | 8 |
| 53 | C, HP, HS, D | Innovating | Local | 9 | 114 | C, HP, HS, D | Complementing | Global | 8 |
| 54 | C, HP | Innovating | Local | 7 | 115 | C, HP, HS | Complementing | Global | 7 |
| 55 | C, HP | Innovating | Global | 8 | 116 | HP, HS | Supporting | Global | 5 |
| 56 | CHP | Complementing | Global | 5 | 117 | HP, HS | Complementing | Local | 5 |
| 57 | C, HP | Innovating | Global | 8 | 118 | C, HP | Supporting | Global | 5 |
| 1 | C, HP, HS, D | Innovating | Local | 9 | 3 | C, HP, HS | Complementing | Global | 7 |
| 58 | C, HP, HS, D | Innovating | Global | 10 | 119 | HS | Complementing | Global | 8 |
| 59 | HS, D | Innovating | Global | 8 | 120 | HP, HS | Supporting | Global | 5 |
| **60** | C, HP, HS, D | Innovating | Global | 10 | **121** | HP, HS | Supporting | Global | 5 |
| **61** | C, HP, HS, D | Innovating | Local | 9 | **122** | HP, HS | Supporting | Global | 5 |
| **62** | C, HP, HS, D | Innovating | Global | 10 | **123** | HP, HS, D | Innovating | Local | 8 |
| **63** | HP | Innovating | Local | 6 | **124** | C, HP, HS | Innovating | Global | 9 |
| **64** | HP | Complementing | Global | 5 | **125** | HP, HS, D | Complementing | Global | 7 |
| **65** | HP | Complementing | Global | 5 | **126** | HP, HS, D | Supporting | Local | 5 |
| **66** | C, D | Innovating | Local | 7 | **127** | C, HP | Innovating | Global | 8 |
| **67** | C, HP, HS, D | Innovating | Local | 9 | **128** | C, HP, HS, D | Supporting | Local | 6 |
| **68** | C, HP, HS, D | Innovating | Local | 9 | **129** | C, HP, HS | Complementing | Global | 7 |
| **69** | C, D | Innovating | Local | 7 | **130** | HP, HS | Complementing | Local | 5 |
| **70** | C, HP, HS, D | Innovating | Global | 10 | **131** | HP, HS, D | Supporting | Global | 5 |
| **71** | C, HP | Innovating | Global | 8 | **132** | C, HP | Complementing | Global | 5 |

In Table 4 we cross-classify the use of technology and the type of technology. As an example, 24 articles describe the use of artificial-intelligence tools for the diagnosis of COVID-19, while 34 the use of telehealth/telemedicine for treatment purposes. All included articles and related analyses are reported in Supplementary material.

**Table 4.**
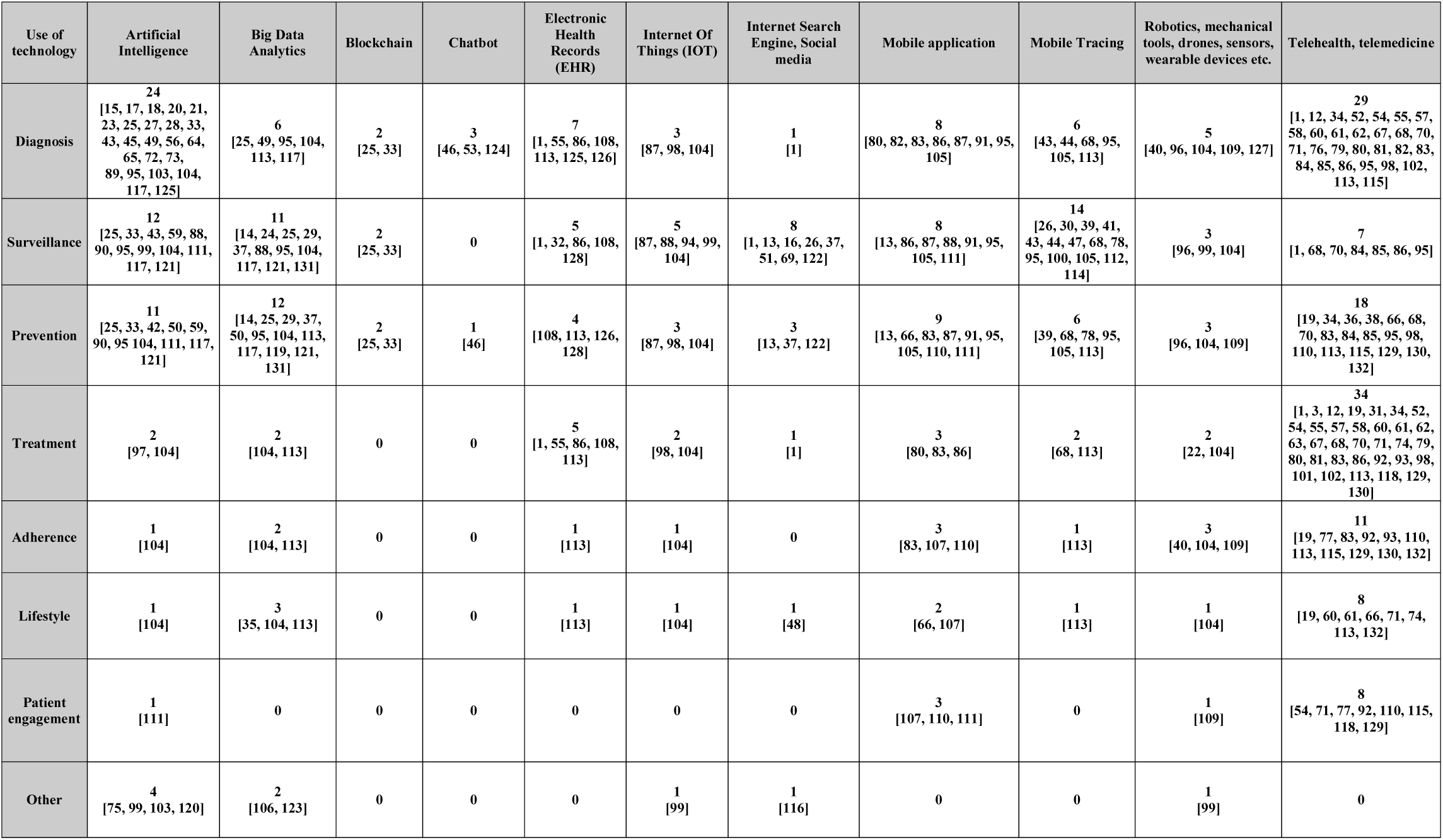
Cross-classification of use of technology (i.e. healthcare need) and type of technology. The first column reports the categories of health needs addressed by technology type (first line).

| Use of technology | Artificial Intelligence | Big Data Analytics | Blockchain | Chatbot | Electronic Health Records (EHR) | Internet Of Things (IOT) | Internet Search Engine, Social media | Mobile application | Mobile Tracing | Robotics, mechanical tools, drones, sensors, wearable devices etc. | Telehealth, telemedicine |
| --- | --- | --- | --- | --- | --- | --- | --- | --- | --- | --- | --- |
| Diagnosis | 24<br>[15, 17, 18, 20, 21, 23, 25, 27, 28, 33, 43, 45, 49, 56, 64, 65, 72, 73, 89, 95, 103, 104, 117, 125] | 6<br>[25, 49, 95, 104, 113, 117] | 2<br>[25, 33] | 3<br>[46, 53, 124] | 7<br>[1, 55, 86, 108, 113, 125, 126] | 3<br>[87, 98, 104] | 1<br>[1] | 8<br>[80, 82, 83, 86, 87, 91, 95, 105] | 6<br>[43, 44, 68, 95, 105, 113] | 5<br>[40, 96, 104, 109, 127] | 29<br>[1, 12, 34, 52, 54, 55, 57, 58, 60, 61, 62, 67, 68, 70, 71, 76, 79, 80, 81, 82, 83, 84, 85, 86, 95, 98, 102, 113, 115] |
| Surveillance | 12<br>[25, 33, 43, 59, 88, 90, 95, 99, 104, 111, 117, 121] | 11<br>[14, 24, 25, 29, 37, 88, 95, 104, 117, 121, 131] | 2<br>[25, 33] | 0 | 5<br>[1, 32, 86, 108, 128] | 5<br>[87, 88, 94, 99, 104] | 8<br>[1, 13, 16, 26, 37, 51, 69, 122] | 8<br>[13, 86, 87, 88, 91, 95, 105, 111] | 14<br>[26, 30, 39, 41, 43, 44, 47, 68, 78, 95, 100, 105, 112, 114] | 3<br>[96, 99, 104] | 7<br>[1, 68, 70, 84, 85, 86, 95] |
| Prevention | 11<br>[25, 33, 42, 50, 59, 90, 95, 104, 111, 117, 121] | 12<br>[14, 25, 29, 37, 50, 95, 104, 113, 117, 119, 121, 131] | 2<br>[25, 33] | 1<br>[46] | 4<br>[108, 113, 126, 128] | 3<br>[87, 98, 104] | 3<br>[13, 37, 122] | 9<br>[13, 66, 83, 87, 91, 95, 105, 110, 111] | 6<br>[39, 68, 78, 95, 105, 113] | 3<br>[96, 104, 109] | 18<br>[19, 34, 36, 38, 66, 68, 70, 83, 84, 85, 95, 98, 110, 113, 115, 129, 130, 132] |
| Treatment | 2<br>[97, 104] | 2<br>[104, 113] | 0 | 0 | 5<br>[1, 55, 86, 108, 113] | 2<br>[98, 104] | 1<br>[1] | 3<br>[80, 83, 86] | 2<br>[68, 113] | 2<br>[22, 104] | 34<br>[1, 3, 12, 19, 31, 34, 52, 54, 55, 57, 58, 60, 61, 62, 63, 67, 68, 70, 71, 74, 79, 80, 81, 83, 86, 92, 93, 98, 101, 102, 113, 118, 129, 130] |
| Adherence | 1<br>[104] | 2<br>[104, 113] | 0 | 0 | 1<br>[113] | 1<br>[104] | 0 | 3<br>[83, 107, 110] | 1<br>[113] | 3<br>[40, 104, 109] | 11<br>[19, 77, 83, 92, 93, 110, 113, 115, 129, 130, 132] |
| Lifestyle | 1<br>[104] | 3<br>[35, 104, 113] | 0 | 0 | 1<br>[113] | 1<br>[104] | 1<br>[48] | 2<br>[66, 107] | 1<br>[113] | 1<br>[104] | 8<br>[19, 60, 61, 66, 71, 74, 113, 132] |
| Patient engagement | 1<br>[111] | 0 | 0 | 0 | 0 | 0 | 0 | 3<br>[107, 110, 111] | 0 | 1<br>[109] | 8<br>[54, 71, 77, 92, 110, 115, 118, 129] |
| Other | 4<br>[75, 99, 103, 120] | 2<br>[106, 123] | 0 | 0 | 0 | 1<br>[99] | 1<br>[116] | 0 | 0 | 1<br>[99] | 0 |

Considering the CIS score, only 7 articles [14, 17, 30, 31, 42, 43, 103] (Figure 2 and 3 [Supp] Supplementary material) resulted above the threshold. These papers can be deemed as valuable considering their scientific impact to date.

Below we provide a summary of the existing digital solutions for the fight against COVID-19 reported in the scientific literature available to date. In order to do this, we discuss some of the retrieved articles deemed most interesting for each healthcare needs.

### 4.1 Diagnosis

To date, digital solutions and innovative technologies have mainly been proposed for the diagnosis of COVID-19. In particular, within the reviewed articles we identified numerous suggestions on the use of AI-powered tools for the diagnosis and screening of Severe Acute Respiratory Syndrome CoronaVirus 2 (SARS-CoV-2) or COVID-19. Most studies propose the adoption of AI tools based on the use of CTs’ data. For example, Zhou et al. [28] developed and validated an integrated deep-learning framework on chest CT images for auto-detection of novel coronavirus pneumonia (NCP), particularly focusing on differentiating NCP from influenza pneumonia (IP), ensuring prompt implementation of isolation. A diagnostic tool like this can be useful during the pandemic, especially when tests such as nucleic acid test kits are short of supply. Nonetheless, performing CT scans as a screening method presents significant limitations, both considering the risk of radiation exposure and operator or machine-type dependence.[20]

Aside from these studies, many Authors propose COVID-19 AI-powered diagnostic tools not based on CT scans data. Feng et al. [17] developed and validated a diagnosis aid model without CT images for early identification of suspected COVID-19 pneumonia on admission in adult fever patients and made the validated model available via an online triage calculator that needs clinical and serological data (e.g. age, %monocytes, IL-6, etc.). Similarly, Martin et al. [46] proposed a chatbot and a symptom-to-disease digital health assistant that can differentiate more than 20,000 diseases with an accuracy of more than 90%.

A further innovative digital technology proposed to support the diagnosis of COVID-19 is the blockchain (or distributed ledger) technology. In one study [33] the Authors recommend a low cost blockchain and AI-coupled self-testing and tracking systems for COVID-19 and other emerging infectious diseases in low middle income countries (LMIC). They developed a low cost blockchain application (app) that requests a user’s personal identifier before opening pre-testing instructions. Following testing, the user uploads results into the app and the blockchain and AI system enable the transfer to alert the outbreak surveillance.

Another interesting digital tool proposed for the diagnosis and triage of patients is chatbots. Chatbots are applications that provide information through conversation-like interactions with users and can be used for a broad range of purposes in health care (patient triage, clinical decision support for providers, directing patients and staff to appropriate resources, and mental health applications). Espinoza et al. underline how chatbots can help evolve triage and screening processes in a scalable manner, [124] concluding that, with institutions becoming more and more familiar with these tools, chatbots can be repurposed in the future for other public health emergencies, as well as for more standard care uses.

### 4.2 Prevention and surveillance

Our literature review suggests that digital technologies can be useful for COVID-19 diagnosis as well as for implementing prevention and surveillance measures.

In Judson et al.,[53] Authors deploy a Coronavirus Symptom Checker, a digital patient-facing self-triage and self-scheduling tool to address the COVID-19 pandemic, that provides patients with 24-hour access to personalized recommendations, and improve ambulatory surge capacity through self-triage, self-scheduling and avoidance of unnecessary in-person care. The majority of patients involved did not make any further contact with the health-system during the subsequent days.

Another topic of paramount importance in the context of healthcare digitalization is epidemiological surveillance. Our review highlights that prevention and surveillance are often considered together in the scientific literature, given that “prevention of COVID-19” can be intended as “prevention of further spread”, which is mainly done through surveillance.

A study by Ferretti et al. [30] analyzes the key parameters of the COVID-19 epidemic spread to estimate the contribution of different transmission routes and determine requirements for successful case isolation and contact-tracing. The authors concluded that viral spread is too fast to be contained by manual contact tracing. The solution is the implementation of a contact-tracing app which creates a temporary record of proximity events between individuals, and immediately alerts recent close contacts of diagnosed cases and prompts them to self-isolate. An example of successful use of a mobile application for contact tracing is the one that the Chinese Government has implemented in Wuhan, as described by Hua et al. [26] A QR code-screening of people was implemented in the Hubei province to monitor people’s movement, especially on public transportation.

Our literature review suggests that another meaningful way to control the spread of an epidemic is through monitoring/surveillance of internet searches and social media usage. Wang et al. [13] used WeChat, a Chinese social media, to plot daily data on the frequencies of keywords related to SARS-CoV-2. The authors found that the frequencies of several keywords related to COVID-19 behaved abnormally during a period ahead of the outbreak in China and stated that social media can offer a new approach to early detect disease outbreaks. Similarly, the Italian words for “cough” and “fever” have been searched in Google Trends to find useful insights to predict the COVID-19 outbreak in Italy, showing a significant association with hospital admissions or deaths in the two following weeks.[133]

Although its potential is irrefutable, the technology behind surveillance and contact tracing apps raises many concerns, as discussed by Calvo et al., [41] the most obvious one being “surveillance creep”, that is when a surveillance tool developed for a precise goal sticks around even when the crisis is solved. Privacy must be a primary concern for the policy makers and a key challenge for designers and engineers that design the digital tools for epidemic control. As already outlined in a previous work by Carullo,[134] in the EU applications to combat COVID-19 should not process personal data whenever possible. The General Data Protection Regulation (GDPR) dictates the principle of privacy by default, that is “by default, only personal data which are necessary for each specific purpose of the processing are processed”. To be compliant with this principle, a preferable approach is therefore to trace the spread of the virus, and therefore alert users, without collecting any personal data. A promising example that goes in this direction is brought by Yasaka et al. [47] with their open source proof-of-concept app for contact tracing that does not require registration or the divulgation of any private data, such as location. Instead, this tool utilizes an ingenious “checkpoint” system, that allows the users to create a peer-to-peer network of interactions and to know if they have been exposed to any risk of infection; diagnosis of infection can be logged into the app, the data is transferred to a central server but stays anonymous.

In regard to the field of prevention, other important digital technologies proposed in the literature include telemedicine and telehealth. Telemedicine does not always cover emergencies, and, as shown also from Lin et al.,[32] many COVID-19 patients may need to go to the hospital for higher level care. For this purpose, Turer et al. [38] propose using electronic Personal Protective Equipment (ePPE) to protect staff (i.e. preventing healthcare workers infections) and conserve PPE while providing rapid access to emergency care for low risk patients during the COVID-19 pandemic. A similar solution has been proposed by Wittbold et al.[82]

### 4.3 Treatment and adherence

Telemedicine and telehealth technologies are also used to increase patient adherence and for treatment purposes. Torous et al.[19] describe the potential of digital health to increase access and quality of mental health care by exploring the success of telehealth during the present crisis and how technologies like Apps can soon play a larger role. Telehealth is seen as a useful solution to deliver mental health care commonly, [135] and during social distancing and quarantine periods.

As another example, Calton et al. [31] deliver some useful tips on the implementation of telemedicine to deliver specialty-palliative care into the homes of seriously ill patients and their families. The authors state that digital divide must be taken into account. Patients need access to a digital device suited for video conference and to an internet connection. For the elderly or the less prone to technology, it may be necessary to identify a caregiver as a “technological liaison” for the patient. To create a successful treatment telemedicine environment, many critical factors are needed: workforce training, high-quality evidence, digital equity, and patient adherence.

### 4.4 Lifestyle and patient engagement

Fewer scientific contributions address the use of digital technologies for lifestyle empowerment or patient engagement. This is probably due to the current phase of the pandemic that has conditioned scientific research to focus primarily on aspects related to more “acute” healthcare needs. However, some articles are present. For instance, Krukowski et al. [115] dealt with the issue of remote obesity management through telehealth methodologies such as electronic scales (e-scales) to remotely measure weight and to maintain patient engagement towards healthy lifestyles.

## 5. DISCUSSION

While SARS-CoV-2 is causing a pandemic worldwide, it is also favoring the rapid adoption of digital solutions and advanced technology tools in healthcare. On the one hand, physicians and health systems may need to track large populations of patients on a daily basis for surveillance purposes.[4] On the other hand, they may need fast diagnostic tests for COVID-19 screening, in order to reduce the workload and enable patients to get early diagnoses and timely treatments. This is also done with the help of digital technologies, which were already available in different industries before the current crisis. These tools have now been quickly implemented in healthcare due to the pandemic.[99]

In this literature review we describe numerous digital solutions and technologies addressing several healthcare needs. The constantly updated scientific literature is a source of important ideas and suggestions for finding innovative solutions that guarantee patient care during and possibly after the COVID-19 crisis.

In the field of diagnosis, digital solutions that integrate with the traditional methods of clinical, molecular or serological diagnosis, such as AI-based diagnostic algorithms based both on imaging and/or clinical data, seem promising and widely used.

As for surveillance, digital apps have already proven their effectiveness, but problems related to privacy and usability remain. For other healthcare needs, several solutions have been proposed using, for example, telemedicine or telehealth tools. These have long been available, but perhaps this historical moment could actually favor their definitive large-scale adoption.

The fact that the digital technologies proposed in the analyzed scientific literature mainly address the fields of diagnosis, prevention and surveillance probably reflects the emergency phase of the COVID-19 pandemic. As time passes, well known digital tools could be proposed for different purposes and healthcare needs, such as adherence, lifestyle and patient engagement.

In addition to the healthcare needs addressed by digital technologies, our review also sheds the light on the most used digital technology tools. Given the early phase of the pandemic, the technologies that have shown to be more easily and quickly implementable can be also considered as the most scalable ones. Among these we report artificial-intelligence tools for diagnosis, big data analytics and mobile tracing for surveillance and prevention, and telemedicine/telehealth, which has proved to be a transversal tool for diagnosis, prevention and treatment. We advocate that many digital technologies quickly implemented in this emergency phase can also be adopted in the following phases of the pandemic.

However, all of this is easier said than done. In the context of the *“Health Care’s Digital Revolution”* brought to the USA (and worldwide) by the COVID-19 pandemic,[4] while private corporations and education institutions have made a quick transition to remote work and videoconferencing, the healthcare system is still lagging behind in adopting digital solutions. This is mainly due to the fact that clinical workflows and economic incentives have been developed for a face-to-face model of care which, during this pandemic, contributes to the spread of the virus to uninfected patients who are seeking medical care. Other than healthcare policies “history”, there are additional limiting factors to the implementation of tools like telemedicine, including a legal framework that is not yet fully designed to regulate the use of innovative IT systems in healthcare, as well as an inadequate ICT infrastructure and an obsolete reimbursement and payment structure.

Many countries are facing these regulatory issues: the challenges for digital health have become a global issue into the public health response to COVID-19 and future outbreaks. Digital tools such as telemedicine should indeed be integrated into international and national guidelines for public health preparedness, alongside the definition of national regulations and funding frameworks in the context of public health emergencies. In order to switch to new digital-based models of care, increasing digital-expertise of health care professionals and educating the population are fundamental issues. Moreover, by implementing a data-sharing mechanism, digitally collected and stored data will be a precious tool also for epidemiological surveillance, that, as discussed earlier, is fundamental in controlling the epidemic spread. Lastly, in order to describe and assess the impact of digital tools during outbreaks, scientific evaluation frameworks should be defined.[36]

This literature review presents some limitations. First, the research was conducted in a period of epidemiological emergency. This determines a large number of daily publications, which is difficult to keep up to date. As a result, we have been forced to select articles in a reduced time span, potentially missing other studies and including studies yet to be peer-reviewed. Secondly, due to the design of the review, the search could not be fully comprehensive, as it was conducted exclusively on Pubmed and MedrXiv. Finally, the articles and concepts included in this preliminary review certainly need to be afterwards integrated at the end of this international emergency phase.

In conclusion, the COVID-19 pandemic is favoring the implementation of digital solutions at a speed and with an impact never seen before. It is therefore mandatory to keep track of the ideas and solutions proposed today to implement tomorrow’s best practices and models of care, and to be prepared in case of future national and international emergencies. We believe that it is worth taking advantage of the push given by the crisis we are experiencing today to implement at least some of the solutions proposed in the scientific literature, especially in those national health systems which in recent years proved to be particularly resistant to the digital transition.

## Data Availability

N/A

## ACKNOWLEDGMENTS

N/A

## COMPETING INTERESTS

The authors declare that they have no competing interests.

## FUNDING SOURCES

The authors declare that they have not received any specific funding.

## Footnotes

1 https://www.medrxiv.ore/

## Notes

### Competing Interest Statement

The authors have declared no competing interest.

### Funding Statement

No funding received

